# Automated segmentation and quantification of histological liver features for MASH/MASLD scoring

**DOI:** 10.64898/2026.02.13.26346163

**Authors:** Kristin Spirgath, Biru Huang, Yasmine Safraou, Mohamed Dahami, Max Kraftberger, Tim-Rasmus Kiehl, Clara H. F. Stockburger, Christian Bayerl, Jakob Ludwig, Noah Jaitner, Anja A. Kühl, Patrick Asbach, Dominik Geisel, Karl H. Hillebrandt, Rebecca G. Wells, Ingolf Sack, Heiko Tzschätzsch

## Abstract

**Background & Aims:** The increasing global prevalence of metabolic dysfunction-associated steatotic liver disease (MASLD) including metabolic dysfunction-associated steatohepatitis (MASH) creates an urgent need for objective methods of histopathological assessment. Conventional histological approaches are time-consuming and rely on interpreter’s experience. Therefore, the results obtained may suffer from high variability and only offer coarse categorisation. In this study, we propose a fully automated, deep-learning-based pipeline for the segmentation and characterisation of histological liver features for MASH/MASLD assessment.

**Methods:** Segmentation was applied to H&E sections from 45 mice and 44 humans with MASH/MASLD. The method, which we named qHisto (quantitative histology), utilises the nnU-Net framework and quantifies key histological components of the MASH score, including macro- and microvesicular steatosis, fibrosis, inflammation, hepatocellular ballooning and glycogenated nuclei. Additionally, we characterized the tissue using novel features that are inaccessible through manual histology, such as the distribution of fat droplet sizes, aspect ratio of nuclei and heatmaps.

**Results:** qHisto parameters showed strong positive correlations with conventional histology scores (fat area *R*=0.91, inflammation density *R*=0.7, ballooning density *R*=0.49) and also with quantitative magnetic resonance imaging (fat area *vs*. hepatic fat fraction *R*=0.87). Our novel scores showed that deformation of nuclei is driven by large fat droplets rather than the overall amount of fat.

**Conclusions:** A key advantage of our method is spatially resolved, precise histological quantification. These features provide a finely resolved assessment of disease severity than conventional categorical scoring. By automating time-consuming and repetitive readouts, qHisto improves standardisation and reproducibility of MASH/MASLD feature quantification and provides scalable, slide-wide readouts that can support histopathologists and enhance clinical assessment and therapeutic development.

**Impact and Implications:** The proposed method provides an objective, automatic tool for comprehensive, histological liver analysis of MASH/MASLD, which can be extended to other diseases and organs. By offering classic and novel quantitative parameters and scores, our method could support histologists in their daily routines and provide researchers with further insight into steatotic liver diseases.

## 1 Introduction

Metabolic dysfunction-associated steatotic liver disease (MASLD) and its progressive inflammatory-fibrotic stage, metabolic dysfunction-associated steatohepatitis (MASH), constitute a major and growing burden in global health in parallel with the epidemics of obesity, type 2 diabetes mellitus and metabolic syndrome. Meta-analyses suggest that steatotic liver disease affects roughly one quarter of the world’s population, making it the most prevalent chronic liver disease worldwide^[1]^. Progression to an inflammatory phenotype with fibrogenesis substantially elevates the risks of advanced fibrosis, cirrhosis, portal hypertension and hepatocellular carcinoma, and is associated with increased liver-related and all-cause mortality^[2]^. While disease terminology has evolved in recent years to better reflect the metabolic drivers^[3]^, the clinical need remains unchanged: to have accurate, reproducible assessment tools that capture disease activity for diagnosis, risk stratification and therapeutic monitoring.

Despite advances in non-invasive biomarkers and imaging, histopathological assessment of liver biopsies continues to serve as the reference standard for diagnosing and staging disease and for adjudicating treatment response in clinical trials^[4, 5]^. Conventional semiquantitative systems such as the MASLD Activity Score (MAS) developed by the MASH Clinical Research Network (CRN) organize steatosis, lobular inflammation and hepatocellular ballooning into ordinal categories^[4, 5]^. However, these scores compress continuous tissue changes into coarse bins, depend on expert knowledge, and are vulnerable to the sampling error intrinsic to needle biopsies. This can also affect the scoring by using a specifically trained algorithm. Substantial intra- and inter-observer variability has been repeatedly documented, undermining diagnostic consistency, statistical power in longitudinal studies, and limiting the detection of disease progression^[5, 6]^. Moreover, conventional scoring can be time-consuming and does not routinely quantify spatial tissue organization or distributional properties of fat vacuoles.

Recent progress in artificial intelligence (AI) and quantitative image analysis offers a path toward objective, fine-grained and scalable histology. Deep learning (DL)-based semantic segmentation has achieved state-of-the-art performance across numerous biomedical imaging tasks by learning tissue-specific features directly from data, enabling pixel-level quantification at whole-slide scale^[7, 8]^. Such pipelines can also be integrated into pathology tools such as QuPath, which can be used to segment and quantify basic features^[9]^. However, most currently available tools focus on isolated readouts (for example steatosis, fibrosis or cell density) and provide only relatively coarse, feature-specific metrics, rather than detailed, multiparametric tissue maps. In pathology workflows, DL assistance has demonstrably improved consistency and efficiency of slide review, illustrating the potential of algorithmic support for complex histologic tasks^[7]^. At the same time, robust deployment in MASH/MASLD faces domain-specific challenges: stain and scanner variability, domain shift across cohorts, class imbalance for rare lesions, and the need for standardized validation against clinical and imaging endpoints.

To address the gaps in conventional histology (conHisto) and the current DL-based tools, we present qHisto (quantitative histology), a fully automated open-access pipeline for segmentation and quantification of histopathological features for MASH scoring, built on the self-configuring nnU-Net framework^[8]^. qHisto is designed to (I) reproduce key elements of established scoring systems with continuous, objective and reproduceable measurements and (II) extend conventional readouts with novel morphometric and spatial descriptors. In order to accomplish these objectives, it is first necessary to segment the relevant structures. The identified structures are then used to define mathematical descriptions of the MASH scores. Finally, the qHisto-derived metrics were validated with quantitative magnetic resonance imaging (MRI) and conHisto. Furthermore, we demonstrate the diagnostic value of the novel parameters. The qHisto methodology involves the transformation of qualitative impressions into standardised numerical values, with the objective of enhancing precision, reproducibility and interpretability in research and diagnostic workflows. Consequently, qHisto has the capacity to facilitate more consistent diagnosis, nuanced disease monitoring and improved evaluation of therapeutic response in MASH/MASLD.

## 2 Methods

### 2.1 Study cohort

#### 2.1.1 Animal cohort

The mouse data used in this study was acquired from the cross-sectional study from Safraou *et al*. in 2025^[10]^. Male C57BL/6l mice (Charles River Laboratories, Sulzfeld, Germany) (*n* = 45; age: 46 ± 7 days; weight: 20.6 ± 2.3 g) were fed a choline-deficient, L-amino-acid-defined, high-fat diet (Diet reference A02082002BR, RESEARCH DIETS, New Brunswick, USA)^[11]^. The animals were assigned to four groups based on diet duration: 12 days (time point = 1, *n* = 10), 21 days (time point = 2, *n* = 10), 81 days (time point = 3, *n* = 9), and 121 days (time point = 4, *n* = 8). An additional control group (time point = 0, day 0, *n* = 8) was maintained on a standard chow diet. Hepatic fat fraction (HFF) was measured by MRI in the mice at each time point. After the imaging procedures, the mice were sacrificed by cervical dislocation while under deep isoflurane anaesthesia, and their livers were collected for histological analysis.

#### 2.1.2 Human cohort

Human patients (*n* = 29) with focal liver lesions (13 male/16 female, age: 65 ± 11, BMI: 26.4 ± 5.4) partially participating in the study of Bayerl *et al*. in 2024^[12]^ were included in our study. In 10 out of the 29 patients’ hepatic fat fraction was measured by MRI 1-3 days before surgery. All patients underwent liver resection, and liver samples were collected during surgery for histological assessment. These 29 subjects were used for the training of the models and the subsequent analysis.

Additionally, 12 histological liver scans from a study from RGW and KH (EA1/055/23, Charité’s Ethics Committee) and 3 scans from the Biospecimen Research Database^[13]^ were exclusively used for model training.

### 2.2 Histology preparation

The liver specimens of the 41 human patients and 45 mice were fixed in 4 % paraformaldehyde (Carl Roth, Karlsruhe, Germany) for histopathological examination. Paraffin embedding and sectioning were performed on both murine and human samples using standard protocols, and 1–2 µm thick sections were stained with haematoxylin and eosin (H&E) for routine morphological evaluation^[10, 12]^. The final histological slides for the 29 patients from Bayerl *et al*. and the 45 mice were digitalised by ZeBanC (Charité) on a NanoZoomer C9600-12 (Hamamatsu, Japan, type: fully automated whole-slide scanner, imaging: brightfield, magnification: 40x, acquisition software: NDP.scan 3.4.1) at a resolution of 0.228 µm/pixel. The 12 patients from RGW and KH were scanned on a Leica Microsystems slide scanner DM6B (Leica, Germany, type: fully automated upright microscope, imaging: brightfield, magnification: 20x, aperture: 0.5, temperature: room temperature, camera: compatible scientific complementary metal-oxide-semiconductor camera, acquisition software: Leica LASX 3.10.1.29575) at a resolution of 0.137 µm/pixel. These digitalised slides were used for the conHisto and qHisto methods.

### 2.3 Conventional histology

Histopathological grading of MASH/MASLD progression was conducted by a pathologist (AAK: more than 18 years of experience) semi-quantitatively according to the MASH-CRN scoring system established by Kleiner *et al*.^[4]^. For assessing inter-observer variability, a second independent pathologist (RK) with more than 10 years of experience scored the 29 human patients.

From the MASH score, 8 features (steatosis grade, steatosis location, microvesicular steatosis, fibrosis stage, lobular inflammation, portal inflammation, ballooning and glycogenated nuclei) were implemented in qHisto and are shown in **Table S1**. with their definition according to Kleiner *et al*.^[4]^. The scores derived from qHisto are sufficient to calculate the MAS score.

### 2.4 qHisto

#### 2.4.1 Automatic segmentation of features

To guarantee the highest possible resolution, the high-resolution scans were divided into smaller tiles (see **Fig. 1**.). The tile size was 5,000 × 5,000 pixels (1,140 × 1,140 µm^2^). For the ballooning model, a smaller tile size of 1,000 × 1,000 pixels (228 × 228 µm^2^) was used because this model had only one label which was small enough to be able to reduce image size and therefore decrease the training duration. The tiles were obtained through the utilisation of the OpenSlide (version 3.4.1) library for Python (https://openslide.org/api/python/).

**Figure 1.**
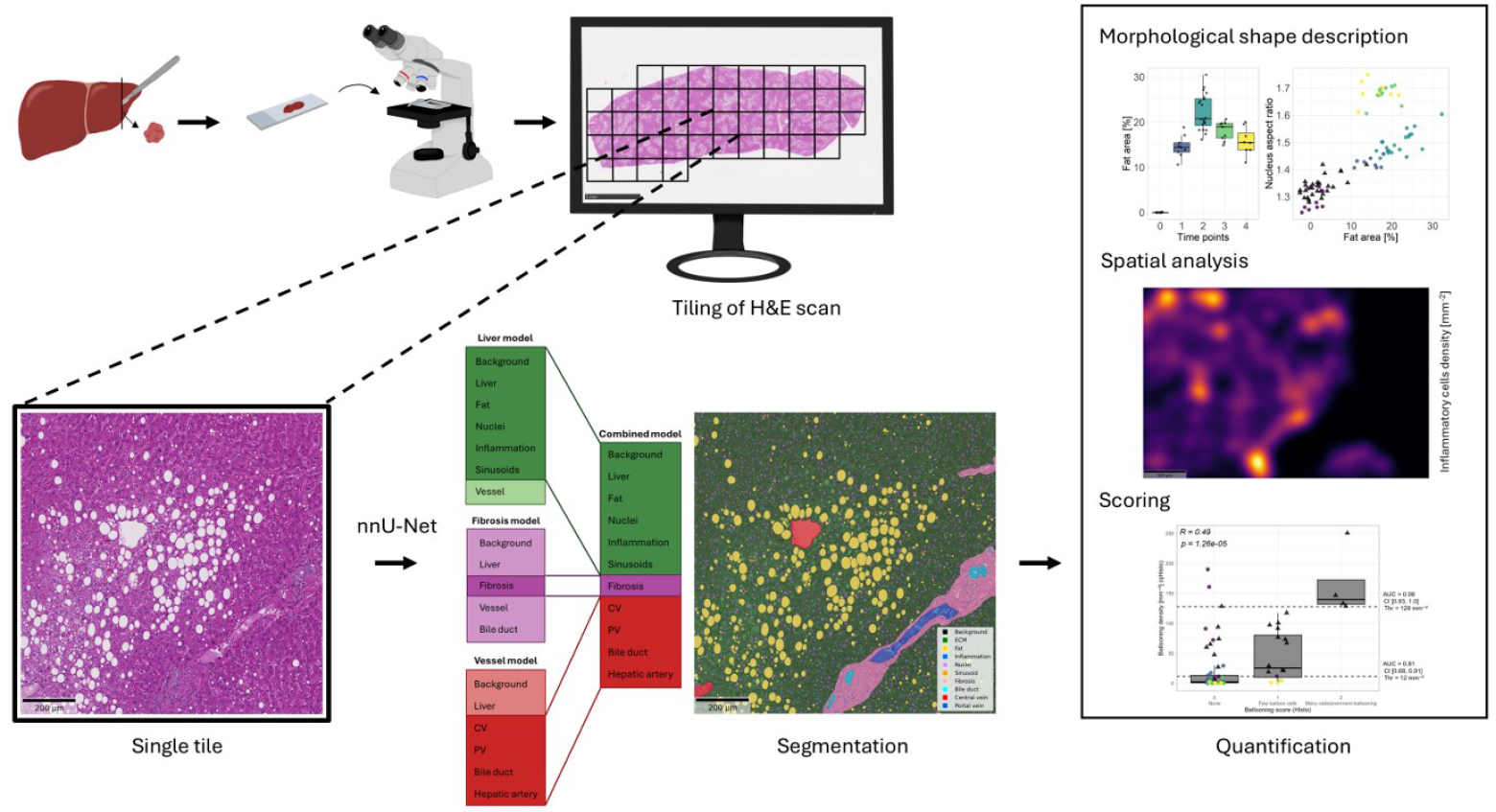
Pipeline of segmentation and quantification algorithm of qHisto. After the preparation of the histological sample, the slide is digitised in high-resolution. The whole scan is divided into smaller tiles, and each tile is then segmented using the trained models. The segmented objects are then quantified using morphological shape descriptors, spatially analysed by extracting heatmaps and histologically scored based on qHisto parameters.

For ground truth, annotations were manually drawn for all relevant features by using the online Computer Vision Annotation Tool (version 2.54.0, https://www.cvat.ai/).

The pipeline consists of five different segmentation models. The first three models were developed during the project, while the last two were trained individually because of possible overlaps in the feature labels. The first model, which was used for steatosis and inflammation, was based on 60 annotated histological tiles that were randomly selected from mice (from Safraou *et al*.^[10]^). These annotations included fat vacuoles, inflammatory infiltrates, hepatocellular nuclei, vessels, sinusoids, liver tissue (parenchyma) and slide background. The second model, which was used to detect fibrosis, comprised 160 manually annotated tiles that were randomly selected from liver tissue of human patients (from Bayerl *et al*.^[12]^). The annotations encompassed fibrotic regions, vessels, bile ducts, liver tissue and slide background. The third model for vessels was trained using 52 annotated tiles, which were randomly selected from mice (19 tiles from Safraou *et al*.^[10]^) and human (33 tiles from Bayerl *et al*.^[12]^). In this instance, a segmentation process was employed to encompass a range of vessel types, including central (CV) and portal veins (PV), hepatic arteries, bile ducts, liver tissue and slide background. A fourth model for ballooned hepatocytes was trained using 1,526 tiles that were randomly selected from human scans (1,151 from Bayerl *et al*.^[12]^, 300 from RGW and KH, and 75 from the database [20]). The fifth model for cells with microvesicular steatosis and glycogenated nuclei was trained on 144 tiles randomly selected from human scans (all from Bayerl *et al*.^[12]^).

Training and prediction of the models were performed on the high-performance computing cluster of Charité and BIH. The automatic tissue segmentation process utilised nnU-Net v2^[8]^ in 2D mode and was configured via the framework’s dynamic task detection, eliminating the need for manual parameter tuning. nnU-Net automatically partitioned the training and validation data using five-fold cross-validation, allocating 80 % of the dataset for training and 20 % for internal validation in each fold. It is important to note that no external test set was defined manually. The evaluation process utilised the framework’s internal ensembling and aggregation of fold-wise validation predictions.

The preprocessing pipeline, incorporating resampling, intensity normalisation, patch size selection, and data augmentation, was managed entirely by nnU-Net’s self-configuring logic. The final models were based on a 2D U-Net architecture with a standard encoder-decoder structure. Accuracy of the prediction was optimized during training using the Dice score per label and averaged over all folds for each model.

Postprocessing was only applied to the vessels due to labels for one structure sometimes being disconnected. To address this, the neighbourhood of each CV and PV label was recursively checked for nearest neighbours within a 250 µm radius. These nearest neighbours were then merged into one structure, and a new centroid was calculated and used for the analysis.

The final performance of the segmentation models was evaluated by calculating the Dice and F1 scores. For objects quantified by the area proportions, the Dice score was of interest (see the bold marking in **Table 1**.). For those objects quantified by number density, the F1 score, where one pixel overlap was defined as a match, was sufficient.

**Table 1.** Quality measures for the different models. Dice and F1 scores for liver, fibrosis and vessel model for the different labels: ECM, fat, inflammation, nuclei, sinusoids, fibrosis, bile duct, CV, hepatic artery, PV, ballooning, microvesicular steatosis and glycogenated nuclei. The relevant score (Dice: if the exact shape/area is of interest; F1 if only the object detection is of interest) for each label is shown in bold.

| Model | Label | Dice | F1 |
| --- | --- | --- | --- |
| Liver | ECM | **0.88** | 0.91 |
|  | Fat | **0.70** | 0.85 |
|  | Inflammation | 0.61 | **0.61** |
|  | Nuclei | **0.74** | 0.74 |
|  | Sinusoids | 0.39 | **0.44** |
| Fibrosis | Fibrosis | **0.81** | 0.90 |
| Vessel | Bile duct | 0.59 | **0.66** |
|  | CV | 0.48 | **0.62** |
|  | Hepatic artery | 0.37 | **0.54** |
|  | PV | 0.58 | **0.62** |
| Ballooning | Balloning | 0.51 | **0.62** |
| Microvesicular steatosis + glycogenated nuclei | ECM | **0.99** | 0.99 |
|  | Microvesicular steatosis | 0.14 | **0.23** |
|  | Glycogenated nuclei | 0.23 | **0.31** |

#### 2.4.2 Quantification of scores and novel metrics

Based on the nnU-Net-generated segmentation masks, qHisto can calculate both conventional scores and novel metrics that are pertinent to MASH scoring. These include the area proportions of segmented features in relation to the full liver area on the slide, which were used for fat and fibrosis. For microvesicular steatosis, ballooning and glycogenated nuclei the number densities in relation to the full liver area were quantified.

For the quantification of the lobular inflammation score by qHisto only inflammatory cells within the liver lobule were considered, and cells segmented within the fibrotic area were therefore discarded. To quantify the lobular inflammation based on the score from Kleiner *et al*. the number of foci normalised to the area of the whole scan was determined using the DBSCAN clustering algorithm from the scikit-learn (version 1.5.1) package. According to the definition of hepatic lobular inflammation by Pai *et al*.^[14]^ in 2019, the minimum number of cells per cluster was set to 2. Additionally, 5 was tested as the minimum number of samples according to Pai *et al*.^[15]^ in 2022. The maximum distance was defined as 80 by optimising the correlation coefficient between the qHisto score and conHisto scores.

Heatmaps were generated for the spatial analysis of the microvesicular steatosis, inflammation, ballooning and glycogenated nuclei. This was achieved by mapping the global centroids of the features to the coordinate space of the image tiles with an empirical bin edge length of 8 µm. The bin counts were converted to densities by dividing by the number of centroids by the tile area. The resulting number density field was smoothed by applying a Gaussian filter and was then rendered as a heatmap. The standard deviation of the Gaussian kernel was set to 10 µm, which empirically provided an effective balance between noise suppression and spatial detail preservation at the cellular scale.

For the morphological analysis of individual hepatocyte nuclei, the aspect ratios were quantified. The aspect ratio was calculated by dividing the length of the largest axis by that of the shortest.

The morphological analysis of the individual fat droplets included the quantification of the diameter and the skewness of the distribution for the diameters for each sample. The skewness was calculated as the standardized third central moment for a variable *x* with observations *x*_1_, …, *x*_*n*_ and sample mean 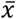 using the following formula:

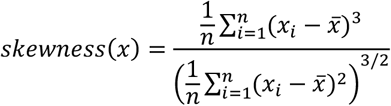

The steatosis location score was determined by analysing the spatial distribution of fat droplets inside the liver lobule^[16]^, using the corresponding score classified by Kleiner *et al*.^[4]^. The zones were defined for each CV. From each CV, a circle with an empirically determined radius of 1.5 mm was defined to estimate the associated PVs for this lobule. A lobule was defined if at least three PVs were found within this circle. The zones were defined as a triangle between neighboured PVs and the CV. These triangles were then subdivided into three equal parts based on the distance between each PV and the CV. Zone 1 is close to the PV and zone 3 is close to the CV, zone 2 lies between those two (see **Fig. S1**.). The number of fat droplets within the designated zones was determined and then normalised to the total area of each zone. The dominant zone was then determined based on the highest density for each feature. A steatosis location score of 0 was assigned if zone 3 was dominant, or if the proportion of zoned fat droplets compared to all fat droplets on the entire slide was less than 5 %. A score of 1 was assigned if the dominant distribution was in zone 1.

A score of 2 was assigned if the distribution was azonal, i.e. if there was no dominant zone or if zone 2 was dominant. A score of 3 was assigned if the distribution was panacinar, i.e. if all zones exhibited a similar distribution and all zones were found to be dominant. This occurred when the percentage of fat droplets in each zone was within 15 % of that in the most affected zone.

To quantify portal inflammation, the total number of inflammatory infiltrates within an empirically determined radius of 400 µm for the humans and 200 µm for the mice around the PV was utilised.

All quantifications were computed using custom Python (Python version 3.10.11) scripts.

#### 2.4.3 qHisto pipeline

As shown in **Fig. 1**., the qHisto pipeline is a fully automated framework for the downstream quantification of histological features. The pipeline starts once a histological liver tissue sample has been digitized. To enable application of the segmentation models, the high-resolution scans are divided into smaller tiles, with the models supporting input sizes of up to 5,000 × 5,000 pixels. Segmented histological features are then quantified for downstream analyses. The full qHisto pipeline will be publicly available under the following repository: https://gitlab.com/Charite-IMI/qhisto_liver.

### 2.5 Hepatic fat fraction by MRI

*Quantitative in vivo* MRI was conducted using a 1.5-Tesla (human) or 3-Tesla (mice) MRI scanner (Magnetom, Siemens, Germany). The protocol comprised clinical T2-weighted imaging as anatomical reference, and a two-point Dixon technique to calculate HFF. All MR images were acquired in a transverse orientation to encompass the entire liver volume^[10]^. Manual drawn regions of interest were used to calculate HFF inside the liver.

### 2.6 Statistical analysis

For the correlation between cardinal values (fat area (qHisto) vs. fat volume and vs. nuclear aspect ratio), the Pearson correlation coefficient was calculated. Concurrently, our results were correlated with scores from conHisto using Spearman’s correlation. For the binary conventional scores, a two sample Wilcoxon t-test was applied.

Translated scores based on qHisto parameters were derived from thresholds by applying an AUC analysis between qHisto and conHisto scores and using the Youden index. The resulting MASH scores were compared between the operator (pathologist A, B and qHisto) by calculating Cohen’s kappa.

## 3 Results

### 3.1 Automatic segmentation of features

The models within the qHisto pipeline were trained to segment multiple different histological features. During the visual inspection of the vessel model, it was observed that CV and PV had been split into multiple labels. Therefore, the post-processing combined multiple labels into one structure. The different models were applied to the scans of human and mice. Example segmentation results from all the models are displayed in **Fig. 2**.

**Figure 2.**
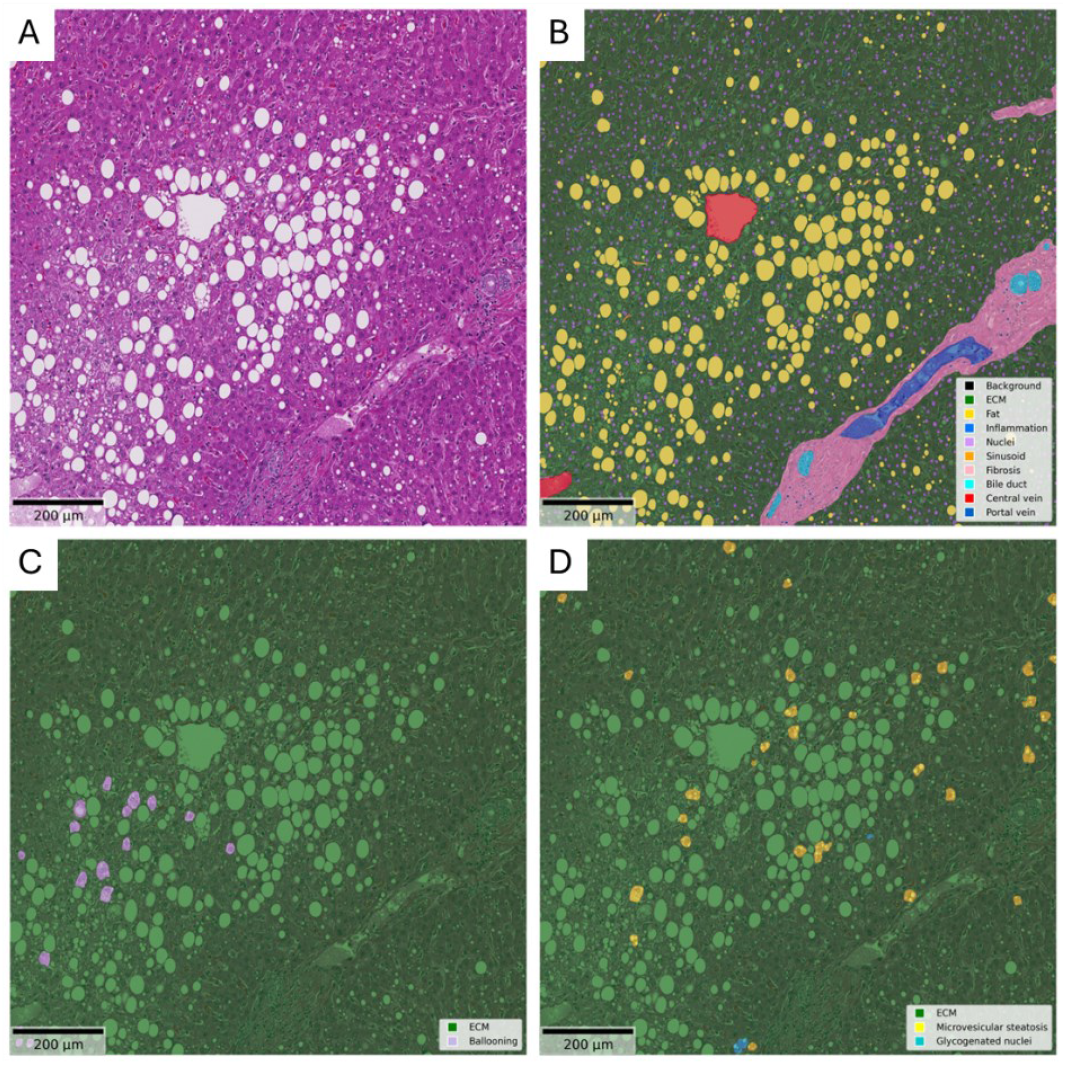
Exemplary segmentation results. (A) 5,000 × 5,000 pixels tile of original human liver scan. (B) Segmentation results in an overlay view with the original scan combined from the liver model, the fibrosis model and the vessel model, (C) from ballooning model and (D) from microvesicular steatosis and glycogenated nuclei model. Please note that, the colours of the labels are semi-transparent and therefore can appear differently in the different areas of the original image.

The performance of the prediction of these features was confirmed by the corresponding Dice and F1 scores averaged over all folds for the different models, as shown in **Table 1**.

### 3.2 Validation of quantified scores

To validate the quantitative histological scores from qHisto, correlations were performed with conHisto and hepatic fat fraction.

For the fat area, the qualitative behaviour over diet and duration was compared with corresponding conHisto and hepatic fat fraction in mice. Similar behaviour was observed for fat area from qHisto and HFF: very low values at time point 0, an increase until time point 2 followed by a decrease until time point 4. In contrast, the scores from conHisto reached a plateau after time point 1 (see **Fig. 3**. (A) to (C)). Quantitative comparison of the fat area from qHisto and the fat volume from MRI revealed a positive correlation (*R* = 0.87, *p* = 0.74×10^-17^, as shown in **Fig. 3**. (D)) in both murine and human subjects. Steatosis grades also exhibited a positive correlation (*R* = 0.91, *p* = 0.67×10^-27^) with the fat area calculated from qHisto when combined with the human data (**Fig. 3**. (E)).

**Figure 3.**
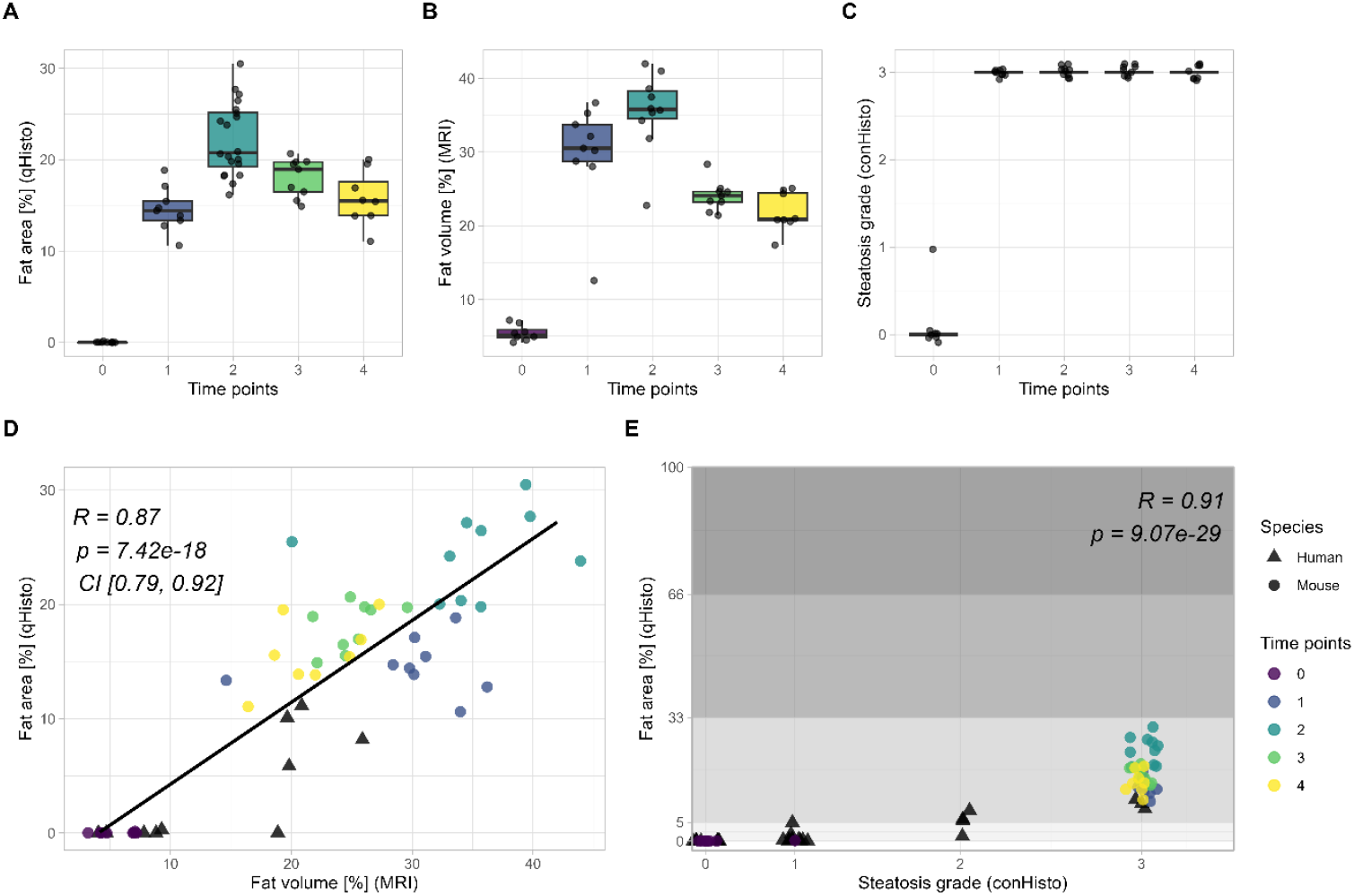
Validation of qHisto scores. (A) Fat area in mice over time by qHisto, (B) fat volume in mice over time by MRI and (C) steatosis score from conHisto. (D) Correlation of fat area (qHisto) and fat volume (MRI) in mice (circle) and human (triangle) (R = 0.87, p = 7.42 × 10-18). (E) Correlation of fat area (qHisto) compared to steatosis score from conHisto for combined human and mice data (R = 0.91, p = 9.07 × 10-29). The grey boxes show the ranges of the steatosis score from conHisto (0: 0 – 5 %, 1: 5 – 33 %, 2: 33 – 66 %, 3: 66 – 100%).

### 3.3 Novel metrics

In addition to the classical shape descriptors, novel metrics were implemented within qHisto to get a deeper insight into changes within tissue.

The nuclear aspect ratio exhibited a positive correlation (*R* = 0.75, *p* = 0.74 ×10^-17^) with fat area in both murine and human subjects (**Fig. 4**. (A)). As it can be seen in **Fig. 4**. (A), time points 3 and 4 form a distinct group.

**Figure 4.**
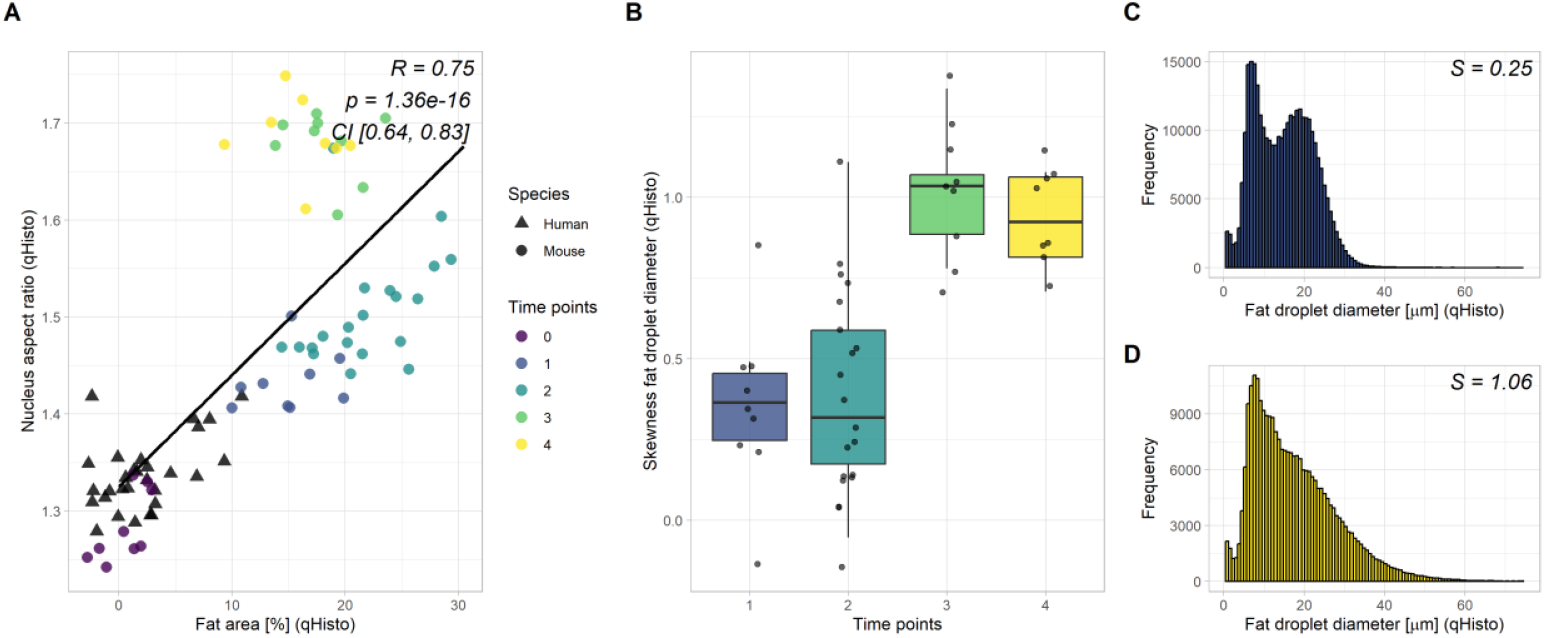
Novel parameters from qHisto. (A) Correlation of total fat area and nuclear aspect ratio in mice (circle) and human (triangle). Normal (solid red line) and deformed (dashed red line) nuclei in human liver tissue. (B) Skewness (S) of fat droplet diameter over time in mice. (C) Distribution of the fat droplet diameter for a mild (S = 0.25, time point 1) and (D) severe (S = 1.06, time point 4) steatosis case in mice.

The skewness of fat droplet diameter also yielded the same distinct group characterised by increased values in mice (**Fig. 4**. (B)). **Fig. 4**. (C) and (D) illustrate two cases of distributions for time points 1 and 4, respectively. This temporal skewness behaviour could not be observed in the human patients, as only one time point was available for each patient.

### 3.4 MASH scoring by qHisto

The parameters quantified by qHisto were used to translate them into MASH scores. These included microvesicular steatosis, fibrosis, lobular inflammation, portal inflammation, ballooning and glycogenated nuclei. The following paragraph compares these quantitative values with the corresponding conHisto scores (see **Table S2**.).

The density of microvesicular steatosis quantified by qHisto did not differ between scores of 0 and 1 from conHisto (corresponding AUC = 0.53; see **Fig. 5**. (A)). There was a positive correlation between the quantified fibrosis area and the score from conHisto (*R* = 0.51, *p* = 0.47 ×10^-5^), and the AUC analysis indicated good separation between the scores (0 *vs*. 1-2: AUC = 0.80, 0-1 *vs*. 2: AUC = 0.83; **Fig. 5**. (B)). Two different minimum sample sizes were tested for the inflammation score. Compared to the conHisto score the qHisto score with a minimum number of inflammatory cells of 2 resulted in a correlation coefficient of 0.55 (*p* = 0.54 × 10^-6^). In contrast, the qHisto score with 5 minimum inflammatory cells showed a higher correlation with the conHisto score (*R* = 0.7, *p* = 0.77× 10^-11^, **Fig. 5**. (C)). Therefore, the qHisto score was calculated using a minimum of 5 inflammatory cells. The inflammation foci density score derived by qHisto yielded a good separation between the conHisto scores (0 *vs*. 1-3: AUC = 0.92, 0-1 *vs*. 2-3: AUC = 0.85, 0-2 *vs*. 3: AUC = 0.74). The qHisto score for the portal inflammation showed a moderate separation between score 0 and 1 with an AUC of 0.69 (**Fig. 5**. (D)). Ballooning density from qHisto correlated moderately with conHisto score (*R* = 0.49, *p* = 0.13 ×10^-4^; **Fig. 5**. (E)) and showed good separation based on AUC analysis (0 *vs*. 1-2: AUC = 0.81, 0-1 *vs*. 2: AUC = 0.98). The density of glycogenated nuclei showed a good separation between scores of 0 and 1 from conHisto and an AUC of 0.91 (**Fig. 5**. (F)) was observed.

**Figure 5.**
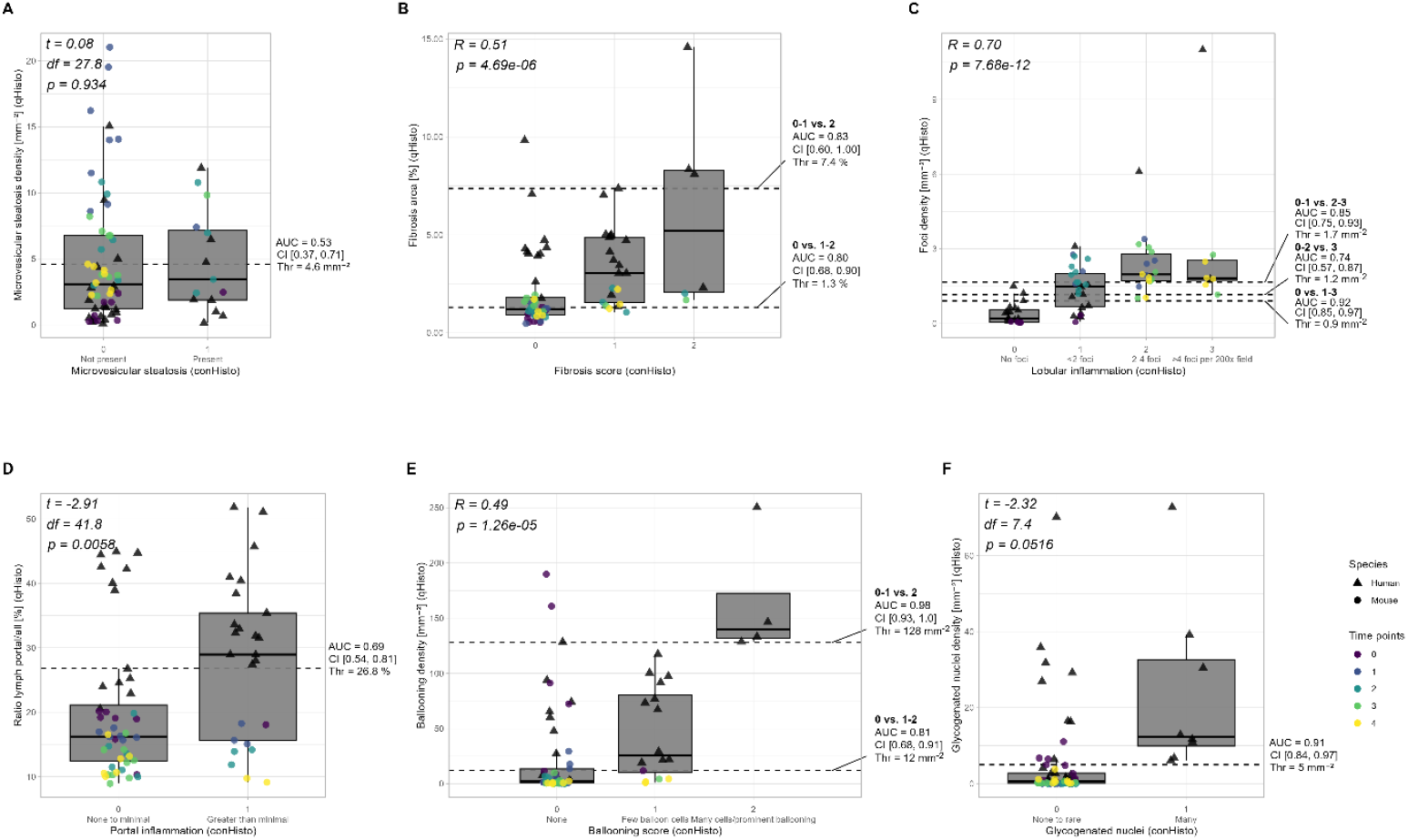
MASH scores by qHisto. Correlations and AUC results in mice (circle) and human (triangle) from (A) microvesicular steatosis (t = 0.08, p = 0.934), (B) fibrosis (R = 0.51, p = 4.69 × 10-6), (C) lobular inflammation (R = 0.7, p = 0.77 × 10-11), (D) portal inflammation (t = −2.91, p = 0.0058), (E) ballooning (R = 0.49, p = 1.26 × 10-5) and (F) glycogenated nuclei (t = −2.32, p = 0.0516). In these plots the scores derived from qHisto are compared to the conHisto scores.

Finally, all scores derived from qHisto were compared to the scores from conHisto from two different experienced pathologists for the patients. The qHisto scores consists of the aforementioned scores and an additional score for the steatosis location and was compared using Cohen’s kappa. This is shown in **Table 2**. Most of the Cohen’s kappa values show a slight or fair agreement between the operators. For fibrosis and lobular inflammation between pathologist B and qHisto and microvesicular steatosis between the pathologists a poor agreement was found. Additionally, some parameters showed moderate agreement between all operators.

**Table 2.** Comparison of inter-operator variability. Cohens Kappa between the different operator (pathologist A, B and qHisto) for the MASH scores derived by qHisto.

| MASH Score | Pathologist A vs. qHisto | Pathologist B vs. qHisto | Pathologist A vs. B |
| --- | --- | --- | --- |
| Steatosis grade | 0.00 | 0.22 | 0.50 |
| Steatosis location | 0.41 | 0.18 | 0.48 |
| Microvesicular steatosis | 0.25 | 0.44 | -0.01 |
| Fibrosis | 0.27 | -0.05 | 0.14 |
| Lobular inflammation | 0.36 | 0.09 | 0.08 |
| Portal inflammation | 0.49 | 0.12 | 0.47 |
| Ballooning | 0.62 | 0.23 | 0.46 |
| Glycogenated nuclei | 0.47 | 0.20 | 0.52 |

### 3.5 Novel spatial analysis

The multiple segmented features enable the possibility of a detailed spatial analysis. In addition to the localisation scores based on existing zone definitions, heatmaps were generated for the microvesicular steatosis, inflammation, ballooning and glycogenated nuclei density. Sample heatmaps are shown in **Fig. 6**., alongside the original scan. It was determined that the cells exhibiting microvesicular steatosis showed clustering but no direct relationship to the vessel structures (see **Fig. 6**. (B)). The inflammation heatmap revealed a homogeneous background and clusters of inflammatory infiltrates in proximity to PVs (see **Fig. 6**. (C)). This heatmap provides an indication of the extent of portal inflammation within the specified tissue (the portal inflammation score in this example was 1 according to classical histology). **Fig. 6**. (D) shows that the ballooning heatmap reveals a heterogeneous background and that ballooned cells are more distant from vessels. Analysis of glycogenated nuclei reveals clustering and avoidance of the area around the CV (**Fig. 6**. (E)).

**Figure 6.**
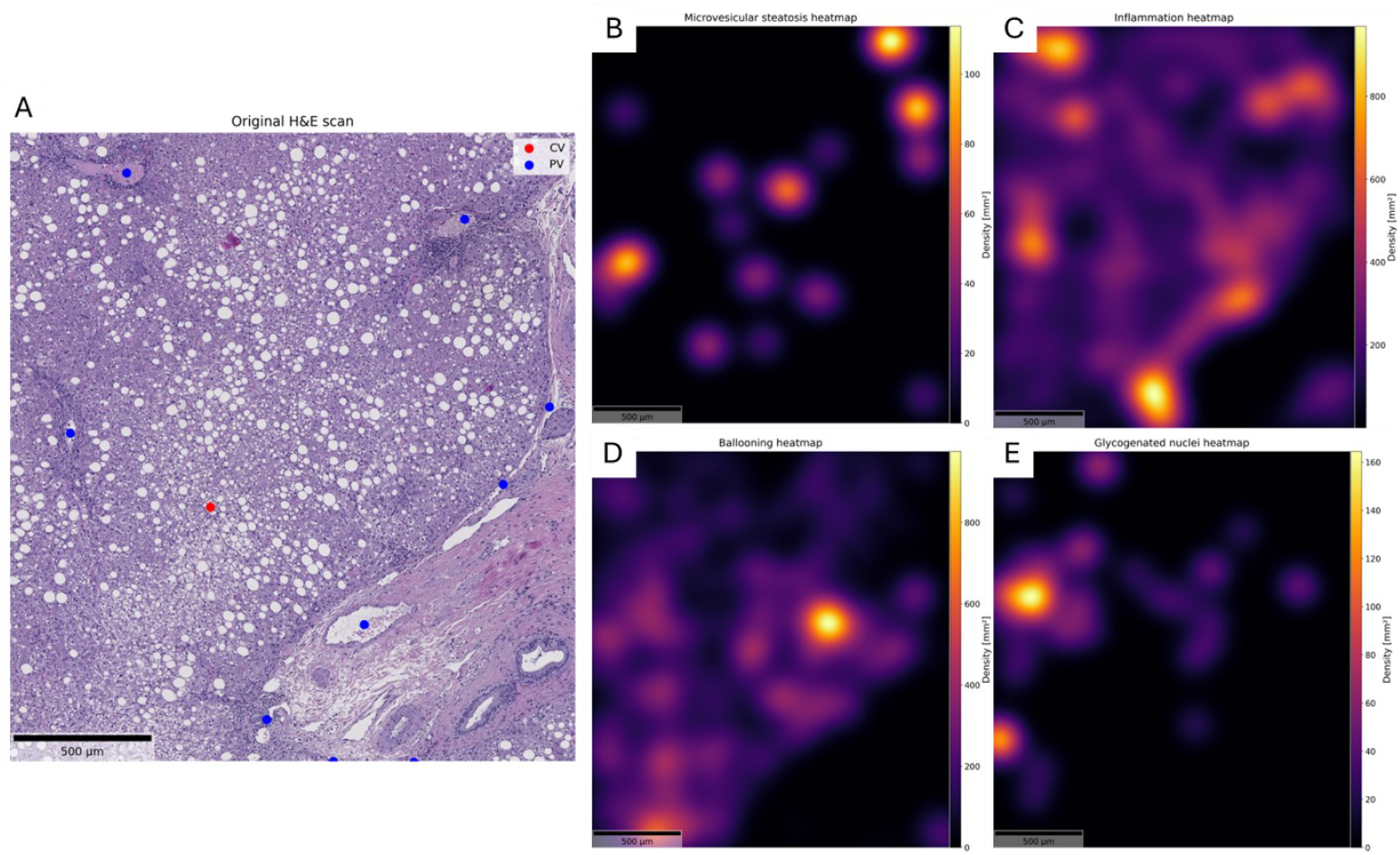
Heatmaps for different histological features. (A) Original human liver scan and corresponding heatmaps for (B) microvesicular steatosis density, (C) inflammation density, (D) ballooning density and (E) glycogenated nuclei density.

## 4 Discussion

This study proposes a framework for automatically segmenting multiple histological features, quantifying derived scores, and generating novel metrics including heatmaps. The algorithms provide deep insights into the tissue composition and can automatically extract detailed information based on the quantified features. Additionally, novel metrics were introduced that give new insights in the tissue’s structure during disease progression. The evaluation and quantification processes produced reliable results comparable to those of conHisto and quantitative MRI.

Other studies that have provided segmentation algorithms based on DL models have been limited in scope by focusing on individual features such as nuclei or steatosis without novel shape descriptors^[17, 18]^. For example, the NucleiSegNet achieved an F1 score of 0.84 for nucleus segmentation whereas qHisto achieved an F1 score of 0.74. However, qHisto not only provides nuclei segmentation but also detailed segmentation of many MASH features simultaneously, showing new possibilities for monitoring changes inside tissue.

Heinemann *et al*. demonstrated the potential for the automated prediction of MASH scores from liver biopsy data using convolutional neural networks, implementing four of these scores^[4, 19]^. While our qHisto pipeline can also estimate eight of the MASH scores, its main strength lies in its detailed morphometric analysis. Cohen’s kappa was higher for the scoring from Heineman *et al*. for the steatosis grade (0.66 *vs*. qHisto: 0.00, 0.22) and fibrosis (0.62 *vs*. qHisto: 0.27, −0.05). The Cohen’s kappa for lobular inflammation (0.24 *vs*. qHisto: 0.36) and ballooning (0.43 *vs*. qHisto: 0.62) were higher when comparing with pathologist A. Unlike approaches optimized primarily to approximate a single score for a slide, qHisto provides continuous, interpretable measurements of individual histological features as well as their spatial distribution within tissue.

QuPath is a widely adopted, interactive platform for digital pathology that provides image viewing, annotation, colour deconvolution and extensible analysis by using scripts and extensions^[9]^. Installation and utilisation of automatic segmentation algorithms such as StarDist can be facilitated within the graphical user interface^[20-22]^. However, QuPath is principally designed for interactive use, and is not optimally suited to large, repetitive, feature-specific computations on whole-cohort scans. In contrast, the qHisto system has been developed to automatically and objectively segment and quantify specific structures in large cohorts, eliminating the need for extensive user interaction.

In recent research, Panzeri *et al*. examined the use of ChatGPT-4 to stage fibrosis in Sirius red-stained liver biopsy images using the MASH-CRN scoring system^[23]^. The study reported strong performance in terms of in-context prompting, achieving almost perfect agreement with a pathologist for pathologist-selected fields of view (Cohen’s kappa: 0.90) and substantial agreement with random crops (Cohen’s kappa: 0.75). By contrast, the Cohen’s kappa for fibrosis with qHisto was 0.27 in comparison to pathologist A. Accurately determining fibrosis stage 4 remained a challenge when scoring with ChatGPT. However, this approach is limited in its capacity to produce pixel-wise quantification and spatial analysis, instead yielding only categorical stage labels. Consequently, large language model-based staging can facilitate initial risk stratification and generate narrative summaries for diagnostic purposes. The qHisto pipeline provides a comprehensive quantitative and spatial analysis of large-scale scan data. Furthermore, it provides reproducible and objective results for the entire scan, eliminating the need for a specific field of view.

It was found to be feasible to reconstruct a significant proportion of the MASH scores within the qHisto pipeline. All parameters derived by qHisto positively correlated with the conHisto score, with the exception of microvesicular steatosis and glycogenated nuclei. However, the separation of glycogenated nuclei based on the AUC remained satisfactory. When comparing the agreement using Cohen’s Kappa, moderate agreement was found between pathologist A and qHisto for ballooning and steatosis location. Fair agreement was found for fibrosis, lobular inflammation, portal inflammation and glycogenated nuclei. There was slight agreement when comparing the scores for microvesicular steatosis between pathologist A and qHisto. The scores for steatosis grade showed no agreement between qHisto and pathologist A and only fair agreement compared to pathologist B. qHisto provides a continuous fat area estimate, while human scoring is only semi-quantitative and therefore Cohen’s kappa can be reduced even though the underlying trend is consistent. The comparison between pathologist A and pathologist B showed that the quality of agreement was within a similar range to the agreement between pathologist A and qHisto. As for the steatosis qHisto can quantify semi-quantitative scores more objective and reproducible and therefore avoid approximations. This could contribute to an improved scoring system.

In addition, the qHisto framework could be applied to both murine and human liver samples using the same pipeline, even though the underlying networks were not consistently trained on both species. Despite these differences in the training data used, comparable quantitative trends were observed in mice and humans, as illustrated in **Fig. 3**. (D) and **Fig. 4**. (A). This cross-species consistency suggests that qHisto captures general, disease-related morphologic patterns, rather than species-specific artifacts, supporting its use in the study of MASH/MASLD in both experimental and clinical cohorts.

Clearly, localisation analysis with pre-defined zones facilitates an objective evaluation of all the extracted features, and not just steatosis. Furthermore, novel parameters were incorporated, including the aspect ratio of hepatocyte nuclei, which is affected by the total fat content. Despite the total fat area decreasing for the third and fourth time points in the mouse study, an increase in the diameter of the fat droplets (as indicated by skewness) was observed. This suggests that the presence of a few large fat droplets causes deformation of the nuclei, rather than the overall fat area. This interpretation is consistent with recent mechanobiology studies demonstrating that fat droplets function as intracellular stressors, deforming hepatocyte nuclei, condensing chromatin, and impairing hepatocyte-specific functions, such as HNF4α expression and albumin secretion^[24]^. To the best of our knowledge, qHisto is the first approach to extend these mechanistic insights by providing a fully automated, quantitative, slide-wide readout of nuclear deformation in routine H&E sections from murine and human MASH/MASLD livers. Overall, our novel parameters can contribute to the development of a new scoring system.

In summary, the qHisto programme can perform a comprehensive and objective analysis of mouse and human liver tissue. This analysis is based on the MASH score, and it provides a detailed evaluation of the tissue through accurate segmentation, as well as quantitative and spatial analysis.

### 4.1 Limitations

The present study has some limitations. Firstly, the fibrosis staging was derived from qHisto using H&E alone, which may explain the relatively modest correlations. Future work will include fibrosis scores based on stains such as Sirius red. However, the exclusive use of H&E highlights the applicability of the framework to the most widely available stain.

Secondly, the size and composition of the training and validation cohorts were limited. These cohorts may not fully represent the broader spectrum of MASH/MASLD encountered in routine biopsy cohorts, which could limit the generalisability of the models to other populations and disease phenotypes. Therefore, the exact values of the derived thresholds should be treated with caution until they are validated in additional cohorts.

Thirdly, Dice and F1 scores were lower for rare or structurally complex features, such as sinusoids, vessels and microvesicular steatosis and glycogenated nuclei. Our models could be improved with additional training data and label refinement, and they could serve as checkpoints for future transfer learning approaches.

## 5 Conclusion

qHisto, a fully automated segmentation and quantification tool for histological scoring of MASH/MASLD was introduced to (I) objective and efficient assessment of MASH features and the corresponding MASH scores. The software can segment and quantify the most relevant features, and is robust for different histological image data, regardless of species, staining or image quality. (II) In addition to the conventional parameters, the qHisto methodology can quantify features that extend beyond the MASH score. These include hepatocyte nuclear shape and spatial analysis using heatmaps. There is potential for qHisto to be expanded in future to include other tissues, features and stainings.

The measures quantified by qHisto were validated by conHisto or quantitative MRI. All parameters demonstrated a positive or even high degree of concordance with the MASH scores, and differentiation based on specific threshold values was feasible, resulting in predominantly elevated AUC values. Consequently, qHisto has the potential to facilitate the diagnosis and advancement of therapeutic interventions for liver diseases such as MASH/MASLD.

## Acknowledgements

The authors acknowledge the Scientific Computing of the IT Division at the *Charité - Universitätsmedizin Berlin* for providing computational resources that have contributed to the research results reported in this paper. URL: https://www.charite.de/en/research/research_support_services/research_infrastructure/science_it/#c30646061

## Data availability statement

The corresponding author will make the data available upon reasonable request. The source code underlying this study will be made publicly available as open source in a dedicated repository in the near future under the following repository: https://gitlab.com/Charite-IMI/qhisto_liver.

## Abbreviations

AUC: Area under the curve
CRN: Clinical Research Network
CV: Central vein
conHisto: Conventional histology
DL: Deep Learning
H&E: Hematoxylin & Eosin
HFF: Hepatic fat fraction
MASLD: Metabolic dysfunction-associated liver disease
MASH: Metabolic dysfunction-associated steatohepatitis
MAS: MASLD activity score
MRE: Magnetic resonance elastography
MRI: Magnetic resonance imaging
PV: Portal vein
qHisto: Quantitative histology

