## Supplement for "Automated segmentation and quantification of histological liver features for MASH/MASLD scoring"

### Title

### Figures

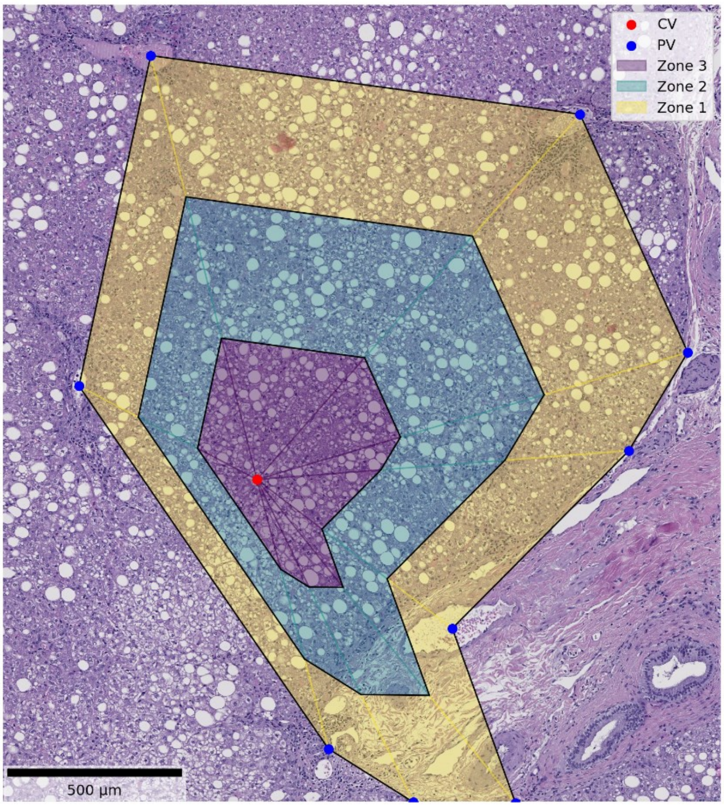


*Figure S1* ***Example of zone definition of a liver lobule.*** *Zone definition for the estimation of the steatosis location score from Kleiner et al.[1] in an exemplary region of a human patient. Zone 3 (in purple) is defined close to CV (red dot), while zone 1 (in yellow) is close to PV (blue dot). Zone 2 (in blue) is defined between the two other zones.*

### Tables

*Table S1* ***MASH scores.*** *Definition of 8 from 14 MASH scores based on Kleiner et al.[1] used in this manuscript.*

| Feature | Score name | Score | Definition |
|---|---|---|---|
| Steatosis | Grade | 0<br>1<br>2<br>3 | < 5%<br>5 – 33 %<br>33 – 66 %<br>> 66% |
| | Location | | |

| | | | |
|---|---|---|---|
| | | 0<br>1<br>2<br>3 | Zone 3<br>Zone 1<br>Azonal<br>Panacinar |
| | Microvesicular steatosis | 0<br>1 | Not present<br>Present |
| Fibrosis | Stage | 0<br>1<br>1A<br>1B<br>1C<br>2<br>3<br>4 | None<br>Perisinusoidal or periportal<br>Mild, zone 3, perisinusoidal<br>Moderate, zone 3, perisinusoidal<br>Portal/periportal<br>Perisinusoidal and portal/periportal<br>Bridging fibrosis<br>Cirrhosis |
| Inflammation | Lobular inflammation | 0<br>1<br>2<br>3 | No foci<br>< 2 foci per 200x field<br>2 – 4 foci per 200x field<br>> 4 foci per 200x field |
| | Portal inflammation | 0<br>1 | None to minimal<br>Greater than minimal |
| Liver cell injury | Ballooning | 0<br>1<br>2 | None<br>Few balloon cells<br>Many cells/prominent ballooning |
| Other findings | Glycogenated nuclei | 0<br>1 | None to rare<br>Many |

*Table S2* ***Statistical results for qHisto parameters****. Thresholds, AUC, R/t and p values for different qHisto parameters compared to conHisto score from human and mice.*

| MASH Score | Range | qHisto parameter | Threshold | AUC | *R/t* | *p[ya1]* |
|---|---|---|---|---|---|---|

| | | | | | | |
|---|---|---|---|---|---|---|
| Microvesicular steatosis | 0 vs. 1 | Density [$mm^{-2}$] | 4.6 | 0.53 | 0.08 | 0.93 |
| Fibrosis | 0 vs. 1-4 | Total area [%] | 1.3 | 0.80 | 0.51 | $0.47\times10^{-5}$ |
| | 0-1 vs. 2-4 | | 7.4 | 0.83 | | |
| Lobular inflammation | 0 vs. 1-3 | Foci density [$mm^{-2}$] | 0.9 | 0.92 | 0.7 | $0.77\times10^{-11}$ |
| | 0-1 vs. 2-3 | | 1.7 | 0.85 | | |
| | 0-2 vs. 3 | | 1.2 | 0.74 | | |
| Portal inflammation | 0 vs. 1 | Inflammation portal/all [%] | 26.8 | 0.69 | -2.91 | $0.58\times10^{-2}$ |
| Ballooning | 0 vs. 1-2 | Density [$mm^{-2}$] | 12 | 0.81 | 0.49 | $0.13\times10^{-4}$ |
| | 0-1 vs. 2 | | 128 | 0.98 | | |
| Glycogenated nuclei | 0 vs. 1 | Density [$mm^{-2}$] | 5 | 0.91 | -2.32 | $0.52\times10^{-1}$ |
